# Early screening for mild cognitive impairment among adults aged 60 and above: A pilot study at a secondary hospital in Ho Chi Minh City, Vietnam

**DOI:** 10.1101/2025.07.10.25331273

**Authors:** Khai Quang Nguyen, The Ha Ngoc Than, Kien Gia To

## Abstract

**Background:** This pilot study explored the feasibility of early screening for mild cognitive impairment (MCI) among adults aged 60 and above at a secondary-level public hospital in Vietnam, with the goal of assessing tool acceptability and implementation potential in routine care.

**Methods:** Among 113 individuals approached, 101 provided consent and 99 completed the screening. Thirty participants met the eligibility criteria and were assessed using six validated tools: the Mini-Mental State Examination (MMSE), Montreal Cognitive Assessment (MoCA), Geriatric Depression Scale-15 (GDS-15), Katz Index of Independence in Activities of Daily Living (Katz Index), Lawton Instrumental Activities of Daily Living Scale (Lawton Scale), and Senior Fitness Test.

**Results:** The enrollment rate was 89.4%, with a screening completion rate of 98%. The MMSE and MoCA demonstrated acceptable internal consistency (Cronbach’s α=0.72 and 0.68, respectively). The GDS-15 (α=0.71) was reliable for depression screening, while the Lawton Scale (α=0.73) effectively assessed functional independence in instrumental activities. However, the Katz ADL showed a ceiling effect, limiting its sensitivity to early impairments.

**Conclusions:** The study confirmed the acceptability of cognitive screening procedures and their practical application in a secondary hospital setting. These preliminary findings provide a foundation for future studies on scalability and integration into routine health services.

## 1. Introduction

Mild cognitive impairment (MCI) is recognized as an intermediate stage between normal cognitive ageing and dementia, characterized by objective cognitive deficits that are measurable through standardized assessments [1,2]. While individuals with MCI may remain functionally independent, they face an elevated risk of progressing to dementia, especially when compounded by factors such as advanced age, depression, and low physical activity levels [3].

Despite growing awareness, MCI remains substantially underdiagnosed worldwide. From 2015 to 2019, about 92% of MCI cases in the U.S. were missed, with 99.9% of clinicians and 99.8% of clinics reporting low screening rates [4]. The challenge is more severe in low- and middle-income countries (LMICs), where access to validated cognitive assessment tools is limited, and awareness of cognitive health is generally low [5]. Around 12.7% of MCI patients develop dementia within one year [6], making early identification essential. Given the modest efficacy and potential side effects of pharmacological options [7], early identification becomes even more critical because it allows timely implementation of non-pharmacological interventions such as cognitive training, which may help delay disease progression [8].

The global demographic shift further amplifies the need for effective early detection strategies. By 2050, the population aged 60 and older is expected to reach 2.1 billion, with 80% residing in LMICs [9]. Vietnam exemplifies this trend, with projections indicating that individuals aged ≥60 will account for over 20% of the national population by 2030 [10]. However, research on early MCI identification remains limited in Vietnam. The absence of standardized cognitive screening protocols within the public health system presents a missed opportunity for timely intervention. Secondary-level hospitals, which play a pivotal role in providing routine healthcare for older adults in Vietnam [11], are well positioned to introduce MCI screening. Nonetheless, concerns remain as to whether adding cognitive procedures in high-volume outpatient settings might disrupt routine workflows or affect screening acceptability, particularly in systems that predominantly focus on physical health.

This pilot study aimed to evaluate the feasibility of early MCI screening among individuals aged 60 and above in a public hospital setting in Vietnam, with a focus on the acceptability and internal consistency of screening tools. The findings may inform future efforts to incorporate structured cognitive assessments into routine care for older adults, supporting earlier identification of cognitive decline and contributing to dementia prevention strategies in resource-limited settings.

## 2. Materials and Methods

### 2.1. Study setting and Participants

This pilot study was conducted at Nhân Dân Gia Định Hospital, a secondary-level general hospital located in an urban area of southern Vietnam. According to internal hospital reports, the facility recorded over 1,000,000 outpatient visits in 2024. Among these visits, approximately 30 to 45 percent of outpatient visits involve individuals aged 60 years and older, among whom 20 to 30 percent present with signs of cognitive decline [12].

Participants were included if they were: (i) aged 60+, (ii) diagnosed with MCI based on Petersen’s criteria [1] and the MCI Working Group of the European Consortium on Alzheimer’s Disease [13], including subjective cognitive complaints (by patient or family), objective decline over the past year or MMSE score between 24 and 29, preserved daily functioning, and no prior dementia diagnosis, (iii) able to read or write Vietnamese, (iv) ability to attend hospital visits, (v) access to supervision for daily activities if needed, and (vi) relatively well-controlled chronic conditions (diabetes and hypertension). Participants were excluded if they: (i) had severe visual or hearing impairments affecting cognitive testing, (ii) had acute or uncontrolled conditions contraindicating exercise or contributing to secondary cognitive decline, (iii) participated in other cognitive programs within the past three months, (iv) were taking medications known to affect cognition, (v) had diagnosed psychiatric disorders affecting cognition, (vi) consumed alcohol regularly at or above 60 grams of pure alcohol per day, or (vii) a GDS-15 score of 5 or higher.

### 2.2. Sampling procedures

All team members received structured training to ensure procedural consistency and enhance data reliability. The research team used different ways to find potential participants for the study. First, researchers checked medical records to see if people had memory or thinking issues. Those who qualified were then contacted during their follow-up visits or by phone. Second, researchers talked to older people in waiting areas, gave them basic information about the study, and invited them to take a screening test. Third, clinicians and healthcare workers referred individuals whom they considered potentially eligible based on a predefined set of criteria established to facilitate rapid identification of suitable participants. These included age over 60, cognitive complaints, and absence of a prior dementia diagnosis. These referrals did not constitute diagnostic assessments but served as an initial triage for study invitation.

To broaden outreach, a community-based strategy was implemented. A Google Form (https://docs.google.com/forms/d/e/1FAIpQLSfa1mlsz5retnvvkWeyltFzetZBeQxKL7tUqjrqGcMGS04HJw/viewform) was distributed through communities and professional groups to identify potential participants, who were then contacted and scheduled for on-site eligibility assessment. This strategy enabled indirect engagement with individuals who may not have been in active contact with the hospital system. The form included four concise Yes/No questions designed to preliminarily identify suitable candidates, focusing on age, subjective memory or attention concerns, absence of a prior dementia diagnosis, and the ability to attend screening visits. Those who responded positively to all four items were subsequently contacted and offered an on-site assessment.

The screening process was implemented as part of a pilot study and was integrated into the hospital’s routine clinical workflow for data collection purposes. It was designed to align with usual care practices without functioning as a standalone clinical protocol, ensuring that no disruption occurred to the regular flow of hospital services.

The screening process began with a structured interview conducted to verify the participant’s age and assess their ability to communicate in Vietnamese. Functional literacy was evaluated through simple reading and writing tasks. Participants were asked whether they could attend hospital visits independently or with assistance, and whether they had access to a caregiver or supervisor when needed. The interview also assessed the presence of subjective cognitive complaints reported either by the participant or a family member, and preserved daily functioning.

Following the interview, medical records were reviewed to determine the presence and clinical stability of chronic conditions, specifically hypertension and type 2 diabetes mellitus. Stability was defined as the absence of hospitalizations or major medication adjustments within the preceding three months. Blood pressure readings obtained during clinic visits were required to be below 140/90 mmHg, and the most recent glycated hemoglobin (HbA1c) level was required to be under 7.5%. Medical documentation was also examined to confirm evidence of objective cognitive decline within the past year, previously recorded MMSE scores if available, and the absence of a prior diagnosis of dementia. Participants were then administered the MMSE as part of the diagnostic criteria for MCI [1,13].

Those who fulfilled all inclusion criteria proceeded to the exclusion screening phase. A brief, targeted clinical examination was conducted to identify any medical or physical factors that might interfere with the validity of cognitive testing or constitute grounds for exclusion. This examination included visual and auditory checks to detect uncorrected impairments, as well as general physical observation to assess signs of acute illness or poorly controlled chronic disease. In parallel, a focused review of the participant’s medical history and clinical records was performed, where necessary, to identify recent engagement in cognitive intervention programs, ongoing use of medications known to impair cognition, and the presence of secondary medical causes of cognitive decline. Psychiatric history and patterns of alcohol consumption were also reviewed to assess for exclusionary conditions. Finally, participants completed the GDS-15 to screen for clinically significant depressive symptoms.

### 2.3. Data Collection and Tools

The study utilized six tools to evaluate cognitive function, depression, daily living activities, and physical fitness. Two cognitive screening tools specifically adapted for the Vietnamese population were used alongside four widely used clinical measures. All assessments followed a standardized sequence to ensure consistency. The Mini-Mental State Examination (MMSE) and Geriatric Depression Scale-15 (GDS-15) were administered first for eligibility screening, followed by the Montreal Cognitive Assessment (MoCA), Katz Index of Independence in Activities of Daily Living (Katz ADL), Lawton Instrumental Activities of Daily Living Scale (Lawton Scale), and Senior Fitness Test (SFT).

#### Mini-Mental State Examination (MMSE)

MMSE is a practical tool for evaluating cognitive function in clinical and research settings. It includes 30 items across 11 tasks covering five cognitive domains. The Vietnamese version, published by the Ministry of Health, was used to better reflect the cultural context and adequately detect cognitive decline in older adults. The intraclass correlation coefficients (ICCs) of MMSE were 0.80 to 0.95, with interrater reliability above 0.90 [14,15].

#### Montreal Cognitive Assessment Version 7.1 (MoCA)

The MoCA assesses six cognitive domains through 30 items and 12 tasks, providing a broader evaluation, particularly of executive function, which the MMSE lacks, and visuospatial abilities, where the MMSE includes only one task. The validated Vietnamese version 7.1, provided by MoCA Test Inc. *(Québec, Canada)*, was used in this study [16]. Previous research has reported strong psychometric properties for this version, including a solid reliability of 0.797 and construct validity supported by confirmatory factor analysis [17].

#### Geriatric Depression Scale-15 (GDS-15)

The GDS-15 demonstrates high diagnostic accuracy in detecting depression among older adults, including in secondary care settings [18]. The Vietnamese version was used to assess the psychological status of participants. Comprising 15 yes/no questions about the past week, its binary format simplifies administration and reduces cognitive load. The GDS-15 has been validated for depression screening in older adults, with α of 0.92 and a high correlation with GDS-30 scores (r=0.966, p<0.001) [19].

#### Katz Index of Independence in Activities of Daily Living (Katz ADL)

The Katz ADL measures six basic daily activities including bathing, dressing, toileting, transferring, continence, and feeding. Each activity is scored based on whether the person can do it independently (1 point) or dependently (0 point). The total score ranges from 0 to 6. A score of 6 means full independence, a score between 3 and 5 shows moderate dependence, and a score of 2 or lower indicates severe dependence. The Katz ADL has been validated for measuring functional independence in older adults, with α of 0.838 and an ICC of 0.999 [20]. After cognitive assessment with the MoCA, the Vietnamese Ministry of Health version was used to assess basic ADLs. It takes 5 minutes to complete [21].

#### Lawton Instrumental Activities of Daily Living Scale (Lawton Scale)

The Lawton Scale checks eight important daily activities needed for independent living. These include using a phone, shopping, cooking, cleaning, doing laundry, moving around, managing medications, and handling finance. Each activity has three to five choices that show different levels of independence. The person’s highest level of ability is chosen as their score. The total score ranges from 0 (complete dependence) to 8 (full independence), with higher scores indicating greater independence. It has been included in national studies under the guidance of the Ministry of Health and applied in population-based surveys in Vietnam [22]. It typically takes 10 to 15 minutes to administer [23].

#### Senior Fitness Test (SFT)

The SFT evaluates key dimensions of physical fitness in older adults. It includes the Chair Stand (lower-body strength), Arm Curl (upper-body strength), Two-minute Step (aerobic endurance), and Timed Up-and-Go (agility and balance). The SFT is highly reliable (ICC=0.80-0.98) and validated against other fitness measures (r=0.71-0.84) [24]. To prevent fatigue, it was administered after cognitive and psychological assessments.

### 2.4. Ethics approval of research

The study was approved by the Biomedical Research Ethics Committee of the University of Medicine and Pharmacy at Ho Chi Minh City (reference number: 625/HĐĐĐ-ĐHYD, dated 05 May 2024) and by the Ethics Committee of Nhân Dân Gia Định Hospital (reference number: 65/NDGĐ-HĐĐĐ, dated 17 May 2024). Caregivers’ consent was prioritized for participants with MCI, ensuring ethical compliance and respect for autonomy. Participants and their caregivers were informed, given time to understand, and signed consent forms voluntarily. The study adhered to the ethical principles outlined in the revised 1987 Declaration of Helsinki.

### 2.5. Statistical Analyses

All analyses were performed using SPSS version 22. Descriptive statistics summarized participants’ demographics (age, gender, and education) using means and standard deviations (SD) for continuous variables, and frequencies and percentages for categorical ones. It also recorded MMSE and MoCA administration times to assess efficiency and practicality.

Feasibility was assessed using three metrics: (i) Enrollment Rate, the proportion of individuals enrolled out of those approached; (ii) Screening Completion Rate, the proportion who completed the screening out of those who agreed; and (iii) Tool Completion Rate, the proportion completing each tool. Rates of 80% or higher were acceptable, with 90% or higher considered excellent [25]. Internal consistency reflects how well items measure the same construct, especially in multi-item scales [26]. An α ≥0.70 is generally considered to indicate reliable internal consistency [27]. A p-value ≤0.05 was considered statistically significant.

## 3. Results

### 3.1 Participant characteristics

A total of 113 individuals were approached for participation through both routine clinical contact and an additional digital outreach using a Google Form. From the online submissions, 18 individuals responded, six were identified as potentially participants, and five presented for on-site screening. Although modest in number, this method demonstrated value in reaching individuals not otherwise engaged with formal care pathways. Among all those approached, 12 (11%) declined participation and two (2%) did not complete the assessment. In total, 99 participants completed the full screening procedure, of whom 30 met the eligibility criteria and were included in the final analytic sample. A detailed overview of the recruitment process and participant flow is illustrated in *Figure 1*.

**Figure 1.**
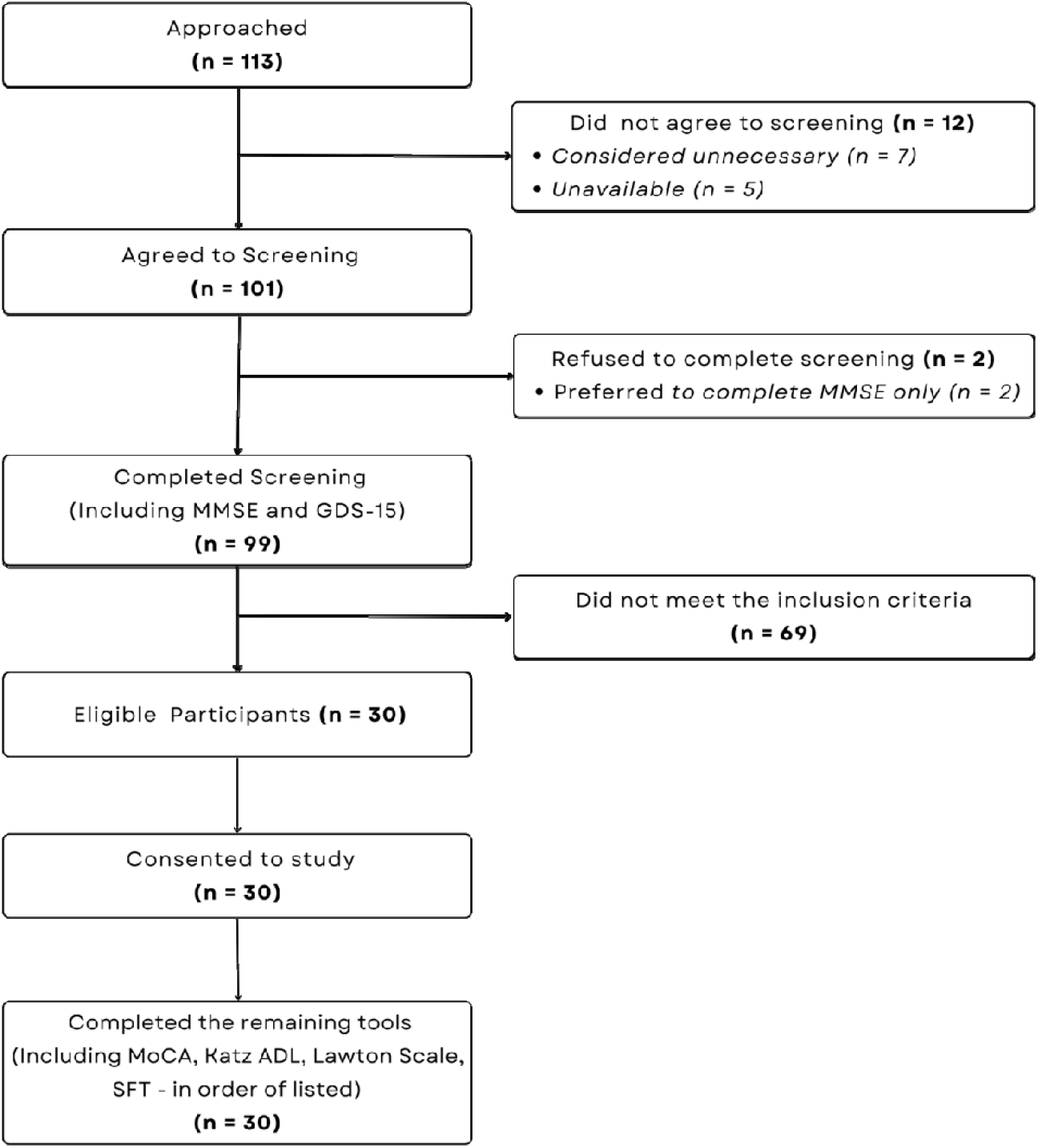
Flow of study’s process.

No significant differences in age, gender, or education were found between individuals who attended the screening and those who were eligible (p>0.05). The mean age of eligible participants was 68.3 years (SD=6). Most (60%) were aged 60–69, and 77% were women. Only 3% had completed primary education. Detailed demographic characteristics are presented in *Table 1*.

**Table 1.** Characteristics of screened participants and those eligible.

|  | Participants<br>attended screening<br>(n=99) |  | Eligible<br>participants<br>(n=30) |  | p-value |
| --- | --- | --- | --- | --- | --- |
|  | n | % | n | % |  |
| **Age group** |  |  |  |  | 0.568* |
| ≤ 59 | 4 | 4.0 | 0 | 0 |  |
| 60-69 | 65 | 65.7 | 18 | 60.0 |  |
| 70-79 | 29 | 29.3 | 12 | 40.0 |  |
| $\geq 80$ | 1 | 1.0 | 0 | 0 | |
| **Gender** |  |  |  |  | 0.833 |
| Women | 79 | 79.8 | 23 | 77.0 |  |
| Men | 20 | 20.2 | 7 | 23.0 |  |
| **Education level** |  |  |  |  | 0.820 |
| Primary School | 3 | 3.0 | 1 | 3.0 |  |
| Secondary School | 28 | 28.3 | 6 | 20.0 |  |
| High School | 40 | 40.4 | 14 | 47.0 |  |
| Intermediate/ College/ University | 28 | 28.3 | 9 | 30.0 |  |
| **Information from screening** |  |  |  |  |  |
| Complained about cognitive problems (Yes) | 96 | 97.0 | 30 | 100 | >0.999 |
| Ensured transportation from home to hospital (Yes) | 80 | 80.8 | 30 | 100 | 0.007 |
| Supervision in daily activities if necessary (Yes) | 86 | 86.9 | 30 | 100 | 0.038 |
| All p-values derived from Fisher's Exact test. |  |  |  |  |  |

### 3.2. Feasibility

The enrollment rate was 89.4% (101/113), and the screening completion rate was 98.0% (99/101). All eligible participants completed the full assessment battery, yielding a 100% completion rate. Average administration times were 10.1 minutes (SD=0.6) for MMSE and 14.7 minutes (SD=0.4) for MoCA. No adverse reaction or safety concerns were reported during the study, indicating that the screening protocol was both feasible and well tolerated.

### 3.3. Performance of Screening Tools

The Vietnamese MMSE showed α of 0.72, indicating solid internal consistency, while the Vietnamese MoCA (version 7.1) had α of 0.68, deemed acceptable. *Table 2* presents α for all instruments used in the study, including MMSE, MoCA, GDS-15, Lawton Scale, and Katz Index. All tools demonstrated satisfactory internal consistency.

**Table 2.**
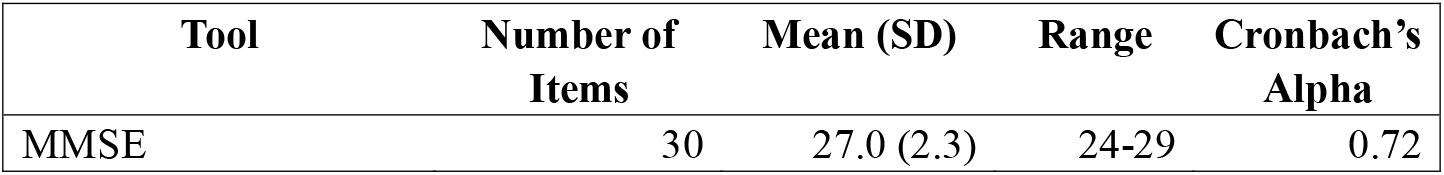
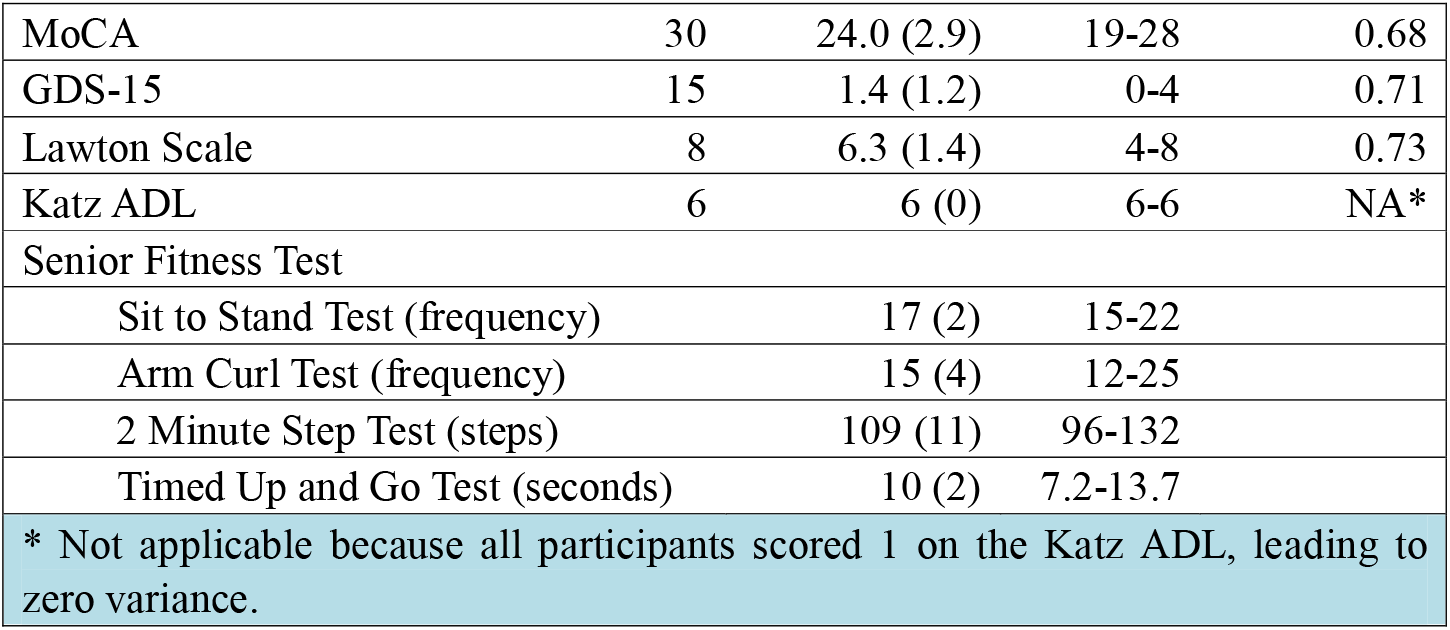
Mean and Standard Deviation (SD) and Internal consistency of MMSE, MoCA, GDS-15, Lawton Scale, Katz ADL, and Senior Fitness Test.

Notably, all participants achieved the maximum score on the Katz Index, revealing a ceiling effect and highlighting the tool’s limited sensitivity in detecting early or subtle functional impairments within this population. The MMSE and MoCA scores were strongly correlated (r=0.723, p<0.001), supporting the concurrent validity of these two cognitive screening instruments (data was not shown).

## 4. Discussion

This pilot study examined the feasibility of implementing early MCI screening among adults aged 60 and above in a secondary-level public hospital in Vietnam. The findings suggest that integrated assessments of cognition, depression, functional independence, and physical fitness were well accepted by participants and demonstrated good internal consistency. While the screening process was not formally embedded in clinical workflows, it demonstrated potential for structured identification of MCI cases and may inform future efforts to explore scalable screening strategies in ageing populations, particularly in LMICs where routine cognitive screening is still limited.

The findings from this pilot study should be interpreted in light of Vietnam’s current healthcare infrastructure, where secondary-level hospitals often serve as the first consistent point of contact for older adults experiencing age-related health concerns [11]. Despite the increasing burden of cognitive impairment [10], routine screening for MCI remains largely absent from public sector practice [28]. The success of this study in achieving high enrollment (89.4%) and screening completion (98.0%) rates within a real-world clinical environment indicates a viable opportunity to embed brief cognitive assessments into routine outpatient visits, especially when aligned with existing workflows. The structured sequencing of tools, beginning with cognitive and psychological measures and ending with physical fitness assessments, may have contributed to reduced fatigue, a known concern in geriatric screening.

Each tool showed adequate internal reliability. The Vietnamese version of the MMSE showed solid internal consistency (α=0.72), consistent with previous studies [14,15], supporting its use in Vietnam’s public health settings. The Vietnamese version of the MoCA (version 7.1) had α of 0.68, likely due to its structure. The MoCA, like the MMSE, has 30 items but covers a broader spectrum of cognitive domains, including executive function and visuospatial skills, through tasks such as drawing, trail-making, and verbal fluency [16]. These tasks require varied cognitive processes, which often lead to lower inter-item correlations and thus reduce internal consistency metrics. Furthermore, internal consistency may be influenced by the heterogeneity of cognitive decline within the target population. As early-stage MCI does not uniformly impair all cognitive domains [29], diverse performance across subdomains may be expected. Prior studies have demonstrated variability in MoCA reliability depending on population characteristics, with α ranging from 0.64 in healthy older adults [30] to 0.76 in populations with a history of psychotropic medication use [31].

In this study, the average administration times for the MMSE and MoCA were 10.1 and 14.7 minutes, respectively. The time required for the MMSE aligns with findings from previous standardized protocols [32,33] and confirms its suitability for use in high-volume outpatient clinics within the Vietnamese public health system. In contrast, the MoCA, though slightly more time-consuming, remained within an acceptable range for clinical application. This duration is slightly shorter than the reported 15 minutes for MCI populations [34] and appears consistent with the cognitive diversity of the sample [35,36]. In cognitively healthy individuals, MoCA has been shown to take as little as 10 minutes [35], while in those with dementia it may exceed 16 minutes [36].

When administration time is considered alongside psychometric properties, the distinct characteristics of each instrument become clear. The MMSE, although brief and efficient, does not comprehensively cover cognitive domains most sensitive to early dementia, particularly executive and visuospatial functions [29]. As such, relying solely on the MMSE may lead to under-identification of early cognitive impairment. On the other hand, the MoCA provides a broader assessment across multiple domains, making it more suitable for detecting subtle and heterogeneous cognitive changes in older adults [37]. Despite a moderate internal consistency (α=0.68), its multidimensional structure remains aligned with the clinical complexity of MCI and preserves its practical value [26]. Since their scores are strongly related (r=0.723, p<0.001), they complement each other in detecting MCI in public hospitals. Given the focus of this study on early screening in older adults, further examination of individual subdomains within the MMSE and MoCA may help refine future protocols. Isolating the most informative components could lead to more efficient and targeted approaches for the early detection of cognitive decline in public health settings.

Beyond cognitive domains, the GDS-15 solid internal consistency with α of 0.71, aiding in evaluating how depression affects cognitive abilities. The Lawton scale (α=0.73) effectively captured early functional decline, while the Katz index exhibited a ceiling effect, making it more suitable for identifying severe impairments [38]. These findings support the use of the Lawton scale for detecting early functional decline, while highlighting the limitations of the Katz index in this context. Building on this, future protocols may consider incorporating complementary tools that assess broader aspects of functional capacity, including elements of social cognition and adaptive behavior, particularly in ageing populations.

The study’s strengths include using validated, culturally adapted cognitive tools for better contextual relevance. The integration of cognitive, depression, and functional assessments provides a comprehensive picture of the participants’ health status, aligning with the multidimensional approach advocated by public health frameworks in ageing populations.

However, the study has limitations. First, the research was conducted in a controlled clinical environment, which enabled more accurate evaluation of the screening tools by minimizing potential confounding variables, such as comorbid physical or psychiatric conditions that could mimic MCI. While this approach enhanced internal validity, it may restrict external generalizability. Second, the sample exhibited a gender imbalance, as 77% of participants were women, likely reflecting greater healthcare engagement among older females. This may also align with demographic realities, as women make up 61.7% of Vietnam’s population aged 60 and above [39]. These potential differences underscore the need for future research with more balanced gender representation. Further, extensive multisite studies are necessary to investigate differences in participant characteristics and screening outcomes across various recruitment channels and healthcare regions.

## 5. Conclusions

This pilot study highlights the potential feasibility of early MCI screening in Vietnam’s public hospital settings. Preliminary results indicate that a screening and multicomponent assessment process can be aligned with routine care without major disruption. Although not formally integrated, the process offered insights into practical implementation and acceptability. Further research is needed to validate standardized protocols and assess their impact in broader clinical settings, particularly in ageing and resource-limited populations.

### Use of AI tools declaration

The authors declare they have not used Generative Artificial Intelligence (AI) tools in the creation of this article.

## Data Availability

All data produced in the present study are available upon reasonable request to the authors

## Acknowledgments

The authors received no specific funding for this work.

The authors would like to thank *Hai Hoang Nguyen, Tan Vo Van*, and *Ly Ngoc Nguyen Tran* for their support, encouragement, and facilitation throughout the research process.

The authors are also deeply grateful to the older adults and caregivers who participated; their trust, time, and openness made this work possible and meaningful beyond research.

